# Artificial Intelligence-informed mobile behavioural interventions to support adolescents’ mental health in schools: protocol for a randomised controlled trial using the MindCraft app

**DOI:** 10.64898/2026.06.17.26355851

**Authors:** Aglaia Freccero, Jack Elkes, Balasundaram Kadirvelu, Ali Versi, Aldo A. Faisal, Lindsay H. Dewa, Martina Di Simplicio, Dasha Nicholls

## Abstract

**Background:** Children and young people (CYP) are particularly affected by mental health problems. Mobile apps provide a scalable and accessible approach to adolescent mental health support, and schools are well-positioned to address multiple risk factors and deliver large-scale interventions. By combining active (self-reported) and passive (sensor-derived) data, mobile apps can model mental states and deliver context-aware support. Artificial Intelligence (AI) enables adaptive, context-aware recommendations tailored to each user. However, there is limited research on AI-based mental health interventions in community CYP. MindCraft is a mobile app designed to monitor adolescents’ mental health using active and passive data and provide AI-informed recommendations (“nudges”). This study aims to investigate the effectiveness of personalised AI nudges delivered through MindCraft on improving mental health outcomes among adolescents in schools in the United Kingdom.

**Methods:** The study is a three-arm RCT using a prospective cohort of secondary school students aged 14-19. Following informed consent, participants complete a baseline online assessment at school and download MindCraft. The primary outcome is the Strengths and Difficulties Questionnaire global and subscale scores. Secondary outcomes include the Eating Disorders Diagnostic Scale, the Sleep Condition Indicator Questionnaire, the Self-Injurious Thoughts and Behaviours Interview, the Self-Efficacy Questionnaire for Children and the World Health Organisation-Five Well-Being Index. Participants are randomised to: (1) an AI-informed intervention group receiving personalised nudges, (2) an active control receiving non-personalised nudges, or (3) a control group with self-monitoring only. Participants use the app for four weeks, with follow-up at one month. Repeated-measures analyses will assess changes across time points.

**Discussion:** We hypothesise that AI nudges will have a greater positive effect on mental health outcomes at one month than general nudges and self-monitoring. Our findings will provide key evidence on the effectiveness of personalised mobile AI recommendations for adolescents’ mental health and inform school-based mental health prevention and early intervention. This study will contribute evidence on the ethical, acceptable, and scalable integration of AI-enabled digital mental health tools within public health and educational systems, with implications for the design of future digital public health interventions and policies supporting their safe integration in schools.

## Introduction

### Background

Children and young people (CYP) are highly susceptible to mental disorders due to developmental changes in emotion, behaviour and cognition, with over 75% of mental disorders emerging before age 25 [1]. The worldwide prevalence of mental disorders in CYP is estimated to be 13.4%, and anxiety, depression, and eating and sleep disorders are prevalent issues among CYP globally [2,3]. Internalising and externalising are two broad categories of behavioural problems, respectively relating to the self and in interaction with the social environment, which in adolescence are associated with an increased likelihood of developing a psychiatric disorder [4]. In the United Kingdom (UK), one in five CYPs is estimated to have a probable mental disorder [5]. However, more than 70% of CYPs struggling do not receive treatment [6], which demands access to cost-effective and evidence-based methods to provide mental health support to CYP at scale.

Given the widespread use of smartphones among CYP [7], digital health interventions are an attractive, cost-effective, and scalable solution for mental health prevention and early intervention among CYP. In the last ten years, there has been a surge in the usage of mental health apps [8]. There are now over 20,000 mental health apps available on the App Store/Android equivalent [9]. Systematic reviews and meta-analyses provide evidence of the positive impact of mental health apps on mental health outcomes [10,11,12], and qualitative studies show that CYP think mental health apps are acceptable [13]. However, most mental health apps are not empirically validated, lack long-term engagement and do not involve CYP in their development, remaining insufficiently tailored to their needs.

Active and passive sensing have become common in digital monitoring technologies for mental health. Active sensors can collect subjective data through repeated self-reports of emotions and behaviours (active data), while passive sensors collect objective data by detecting changes in the physical environment without user participation (passive data), including GPS, accelerometer, microphone and battery usage. Active and passive data serve as digital behavioural markers, enabling modelling of individuals’ mental health states across their contextual and temporal dimensions [14]. Based on this data, Ecological Momentary Interventions (EMI) can deliver personalised behaviour change support in real-time and in natural settings [15].

Artificial Intelligence (AI) and Machine Learning (ML) present new opportunities for real-time interventions. A systematic review of 17 studies examined the feasibility of using AI-enabled mobile solutions using active and passive data to support mental health [16]. Across the 17 studies, the most common use of AI was to predict mental health outcomes, provide natural language support, and develop risk profiles. Only 2 studies used AI/ML to deploy context-sensitive notifications [17] and CBT-based JITADs [18]. All studies were conducted in adults, with small sample sizes and short intervention durations. None of these studies was carried out in the UK. Only 3 studies were RCTs, and they had small sample sizes. Overall, AI/ML can be integrated into mental health apps, but the lack of evidence and weaknesses in study designs highlight the need for high-quality RCTs to demonstrate the effectiveness of AI-based apps in improving mental health.

MindCraft is an AI-based mental health app that combines active and passive tracking to monitor CYP’s emotions and behaviour [19] and has already been tested for feasibility as a mental health risk prediction tool in a non-clinical population of adolescents [20]. A new version of the MindCraft app has been developed in collaboration with CYP, which features a reinforced learning-based recommendation system designed to promote behavioural change and enhance CYP mental health. This system dynamically detects early signs of changes in mental health states from active and passive data collected through the app and responds by delivering personalised AI-informed recommendations (“nudges”). Intervention outcomes are measured in real-time and fed back to generate novel, individually tailored candidate nudges. This technology learns specific CYPs’ emotional and behavioural profiles and contexts and recognises by “trial-and-error” successful from non-successful interventions.

### Research Purpose and Aims

School-based research suggests that the school environment plays a critical role in shaping students’ behaviour and significantly impacts CYP’s health [21]. This makes schools well-placed to identify and address multiple determinants of mental health risk at the individual and community levels. A recent meta-analysis showed a small positive effect of brief school-based interventions on mental health outcomes for up to 1 month, 6 months, and 1 year [22]. In particular, digital mental health intervention programs offered in schools could provide a readily accessible, flexible means of educating, empowering, and supporting adolescents in maintaining good mental health [23]. However, there is a lack of research on the efficacy of digital interventions, particularly those informed by AI/ML, on mental health outcomes in community CYP.

Therefore, this study aims to examine the effectiveness of personalised AI-based recommendations (“nudges”) compared to generic psychoeducation recommendations and self-monitoring on mental health outcomes in CYP within secondary schools across the UK. Outcomes will be assessed using the Strengths and Difficulties Questionnaire (SDQ), a validated tool that captures both internalising (e.g., anxiety, depression) and externalising (e.g., aggression, hyperactivity) behaviours, enabling a comprehensive evaluation of changes in mental health following the intervention. We hypothesise that AI nudges will be more effective than non-personalised digital self-help using generic CBT principles (active control group) and self-monitoring (control group) in reducing SDQ scores at 4 weeks follow-up.

## Methods

### Study Design

The study will be conducted and reported according to the Consolidated Standards of Reporting Trials (CONSORT) and CONSORT-EHEALTH for improving and standardising evaluation reports for mobile interventions [24]. We conducted an external pilot from October to May 2025 to test the quality of the trial design and conduct with respect to recruitment, adherence, outcome assessment, sample size, and follow-up. After the pilot, analyses were conducted to calculate the sample size needed for the main trial. We have also released an improved version of the MindCraft app (e.g., fixed bugs, upgraded systems, and improved the user interface).

The study design is a multicentre interventional 3-arm randomised parallel-group trial. Once trial eligibility has been determined and informed written consent to participate has been obtained, participants will be individually randomised into one of three arms:

1. Experimental: Receipt of personalised AI-based recommendations within the MindCraft app in addition to self-monitoring
2. Active control: Generic psychoeducation recommendations within the app and self-monitoring through the app
3. Control: Access to self-monitoring through the app only

This three-arm design supports attribution of observed effects specifically to the personalised AI component while maintaining ethical access to basic self-monitoring functionality for all participants.

### Recruitment

The study will take place in secondary schools in the UK. The research team will contact school staff via email to provide information about the study. Upon receipt of a reply, a member of the research team will schedule an online meeting to provide an overview of the trial to relevant staff and discuss the school’s participation. We aim to recruit around 5-10 schools, depending on the number of students they accommodate. If the school agrees to participate, a member of the school staff, acting as a liaison for the study, will circulate information about the study and invite students in years 10 to 13 (aged 14-19) to participate. A date for baseline data collection on school premises will be agreed with the school. If preferred, schools will have the option to carry out the study remotely by circulating study information to students and a consent-to-contact form. Interested students will be contacted, and following online consent, baseline and follow-up measures will be collected via an online Qualtrics questionnaire sent to the participant’s email.

All potential participants and their parents/guardians (if under 16) will be provided with copies of the Participant Information Sheet and the Parent/Guardian Information Sheet. Students aged 16 or older will provide informed consent electronically via Qualtrics (https://www.qualtrics.com/uk/) on the day of data collection. For participants under 16 years old, parental or guardian opt-out consent will be used. Parents/guardians will be informed by the school that the study is taking place and will sign and email the Opt-out Consent Form back to the researcher if they do not wish their child to participate. Parents will have at least two weeks to complete the opt-out form before any data is collected from children on school premises. Students under 16 years will also provide written assent via Qualtrics on the day of data collection.

#### Inclusion Criteria

– Young people (aged 14-19) in years 10 to 13 attending any school in the United Kingdom that has been approached
– Sufficient English to allow completion of experimental measures and use of the app
– Access to an iOS or Android-compatible smartphone with embedded activity monitor
– Have the capacity to consent (if over 16 years old)/assent and seek consent from parents/guardians (if under 16 years old)

#### Exclusion Criteria

– Have learning difficulties, organic brain disease, severe neurological impairment that prevents independent use of smartphone app
– Have no access to a personal mobile phone

### Procedure

Participants’ consent/assent is obtained via Qualtrics on the day of baseline data collection. After providing consent, participants will complete a baseline mental health questionnaire (see Outcomes) via Qualtrics on their personal mobile device, separate from the Consent Form. Completing the questionnaire will take approximately 30 minutes, following which participants will be able to download the MindCraft app from the app store.

Participants’ data will be pseudonymised, and each participant will be assigned a unique study ID to input in the questionnaire and access the app. This ID also ensures that each participant receives only their assigned intervention, preventing cross-arm contamination.

Participants will be instructed to record data every day for 4 weeks, and will be followed up after approximately 4 weeks (Table 2), scheduled according to school availability. Individual participants will not be reimbursed. Participating schools will have the opportunity to take part in research and mental health-related activities at Imperial College.

**Table 2.**
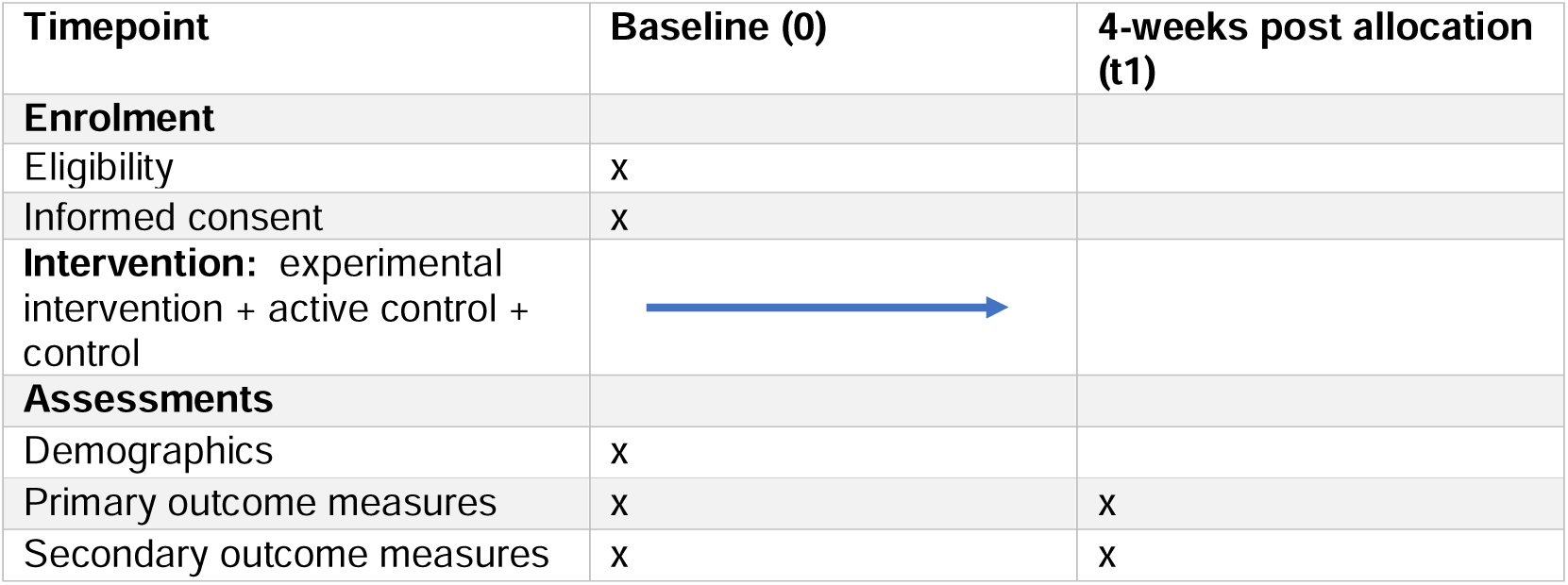
Schedule of enrolment, interventions and assessments.

| Timepoint | Baseline (0) | 4-weeks post allocation (t1) |
| --- | --- | --- |
| **Enrolment** |  |  |
| Eligibility | x |  |
| Informed consent | x |  |
| **Intervention:** experimental intervention + active control + control |  |  |
| **Assessments** |  |  |
| Demographics | x |  |
| Primary outcome measures | x | x |
| Secondary outcome measures | x | x |

Randomisation will be conducted using a secure, computer-based tool to ensure unbiased allocation of participants across the three study arms. Simple randomisation will be used, giving each participant an equal chance of assignment to any arm. Allocation will be implemented through a central web-based system, which conceals the sequence from outcome assessors and trial staff until the moment of assignment. Participants will be allocated to experimental or control interventions on the day of baseline data collection, with the sequence generated by an independent statistician to ensure proper allocation concealment.

Participants in both the intervention and active control groups, as well as the outcome assessor, will be blinded to intervention allocation. The interventions are designed to be similar in appearance and delivery to maintain blinding (see Intervention). Only the trial statistician will have access to the allocation list and will not disclose this information until after trial completion. Unblinding is not expected, and would only occur in exceptional circumstances where knowledge of a participant’s allocation is required for safety.

### Intervention

MindCraft app is available for free download from the Apple App Store and Google Play Store (Figure 1a). The study ID code provided at baseline is required to access the app (Figure 1b). The MindCraft app comprises 3 main tabs in its User Interface: the main tab, the progress tab, and the settings tab (Figure 2). The MindCraft app is designed with a modular, adaptable framework to facilitate future improvements, support different study questions, and provide customisation options to meet individuals’ needs and enhance engagement [19].

**Figure 1.**
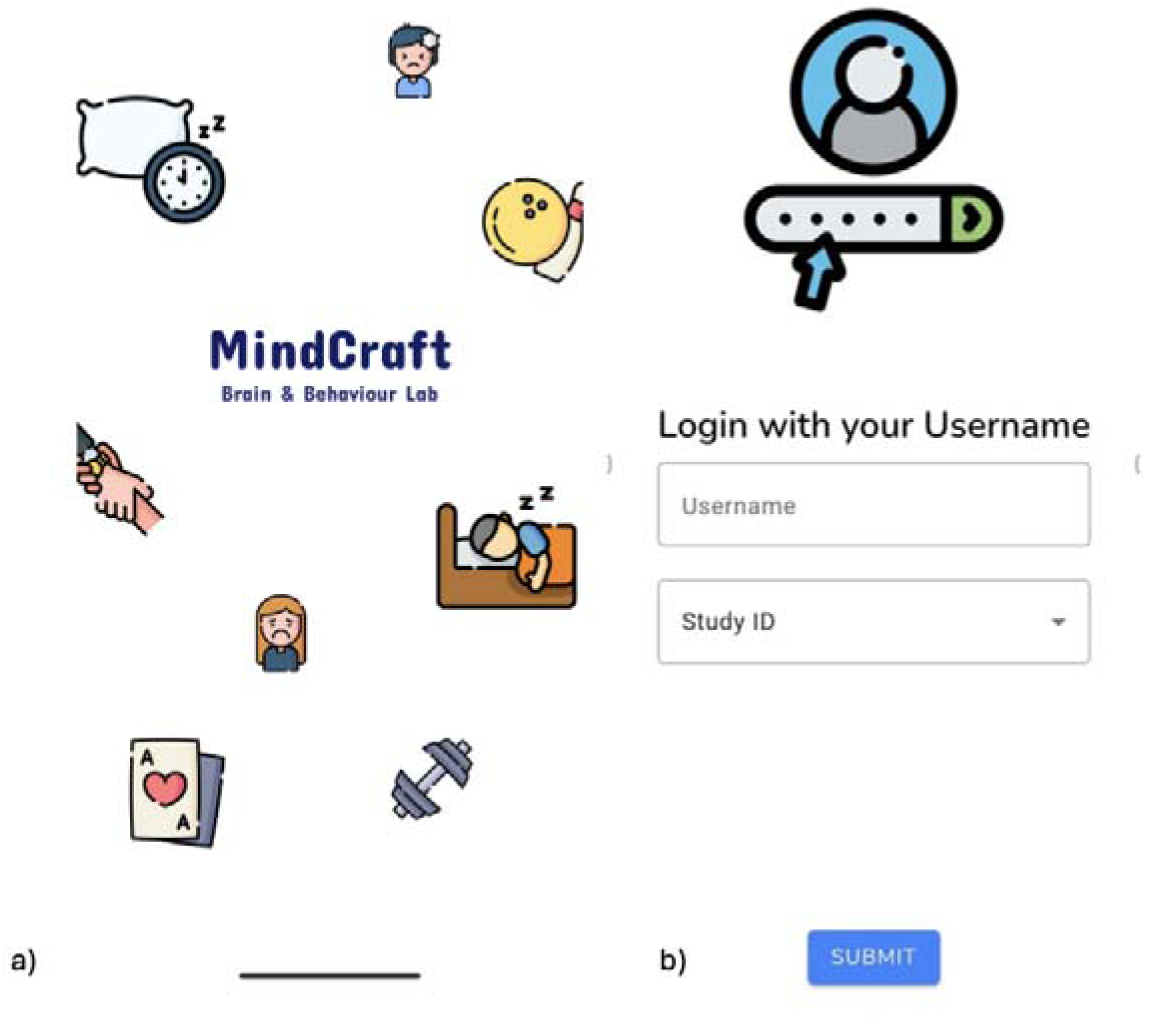
Login screen.

**Figure 2.**
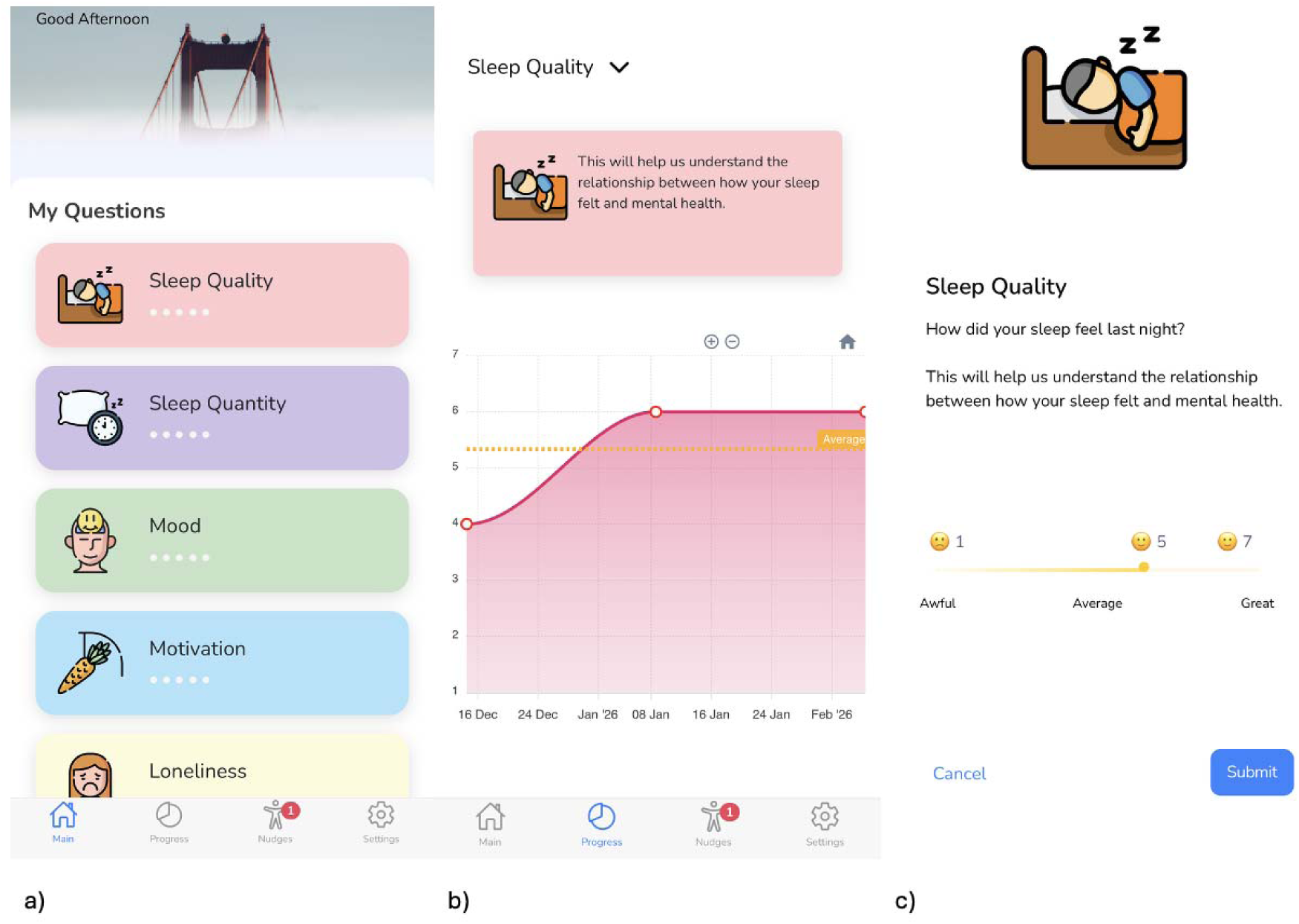
Main Tab and Progress Tab. · <u>Main tab</u> (Figure 2a): The main tab is the first view of the app when launched, where users can self-report a mood and well-being questionnaire. The questionnaire contains 8 required questions (sleep quality, sleep quantity, mood, motivation, anxiety, loneliness, appetite, and exercise), as well as 11 additional questions (racing thoughts, negative thinking, hopefulness, headaches, irritability, confidence, sociability, energy levels, productivity, self-care, leisure, and a custom metric called “your measure”). The user can manage the optional questions through the settings tab. Each question can only be answered once a day and disappears after each submission. Most self-reported questions (such as mood and sleep quality) are on a 1-7 scale and include a slider (Figure 2c). Other questions (such as sleep quantity and exercise time) collect numerical data from text boxes, and the input is type-checked to ensure only valid values are stored (e.g., numbers, not text). · <u>Progress tab</u> (Figure 2b): The progress tab charts the user’s self-reported updates throughout the study. Selecting the data of interest displays the historical data and the average score for the selected period in the graph view.

**Figure 3.**
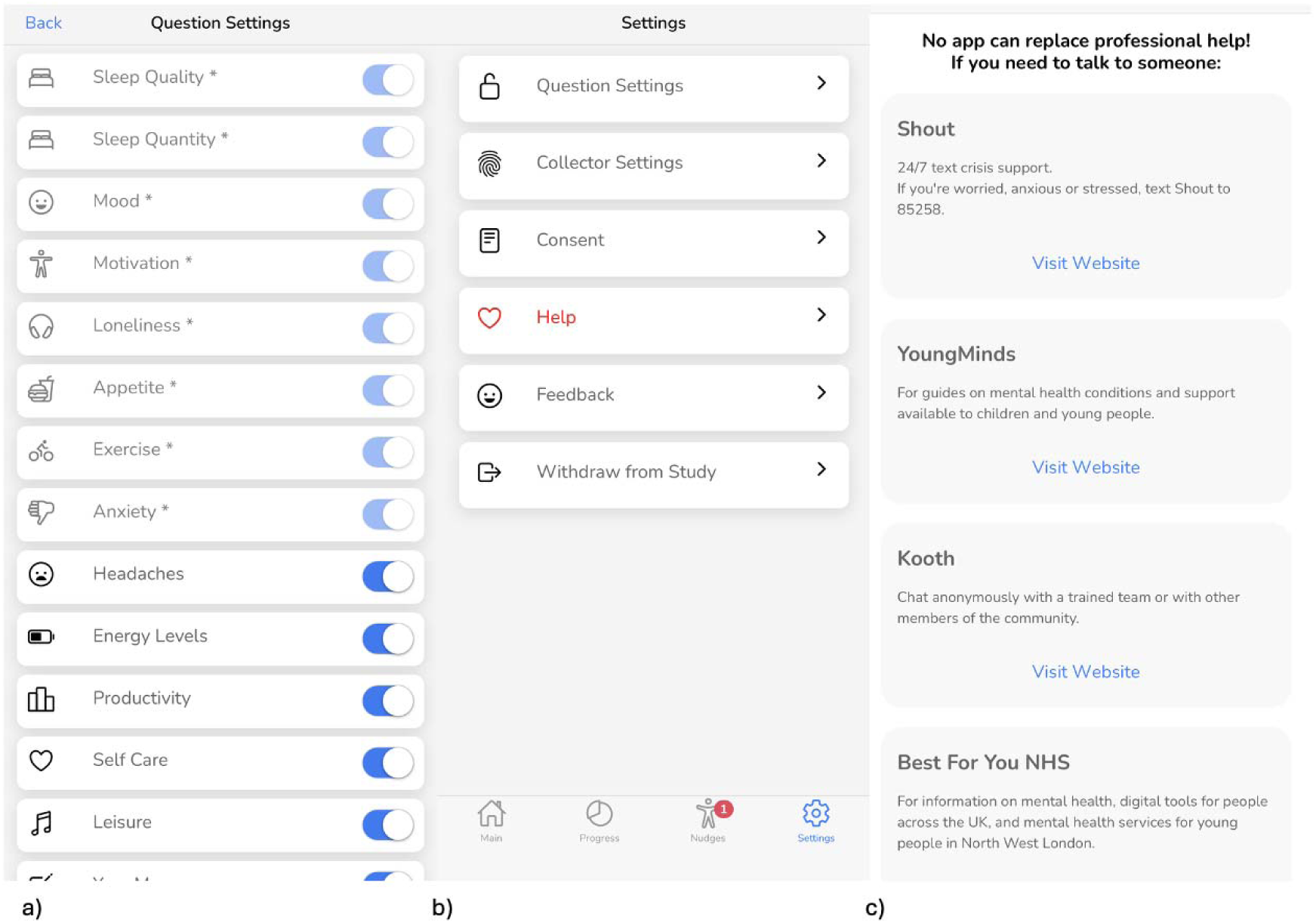
Settings Tab. · <u>Settings tab</u> (Figure 3): In the settings tab, the user can manage the active and passive data settings, read the privacy policy, and withdraw from the study. Users can set their active and passive data-sharing preferences during initial onboarding and modify them at any time in the app’s settings. Participants are free to select among 8 required and 11 optional trackers (Figure 3a). Users can choose which passive data they wish to share, ensuring that they have control over their privacy (Figure 3b). The passive trackers include phone and app usage, battery level, location (collected when there is a significant change in the user’s location), steps, background noise (collected only when the app is in the foreground) and ambient lighting. The “Help” option under the Settings tab allows users to seek support when needed (Figure 3c). To facilitate this, MindCraft lists four external organisations: Shout, YoungMinds, Best For You NHS, and Kooth - along with a brief description of each organisation and links to their websites. · <u>Nudges tab (</u>Figure 4): In the nudges tab, participants in the experimental and active control groups will access 3 potential nudges from a list developed in collaboration with CYP (see PPIE). The content of the nudges draws from cognitive behavioural therapy (CBT) and behavioural activation, as well as mental imagery and mindfulness techniques. These include tips, psychoeducation and exercises. Participants receive a notification when a new selection of nudges becomes available in the nudges tab. Once participants complete the suggested activity, the nudge is considered complete and can be rated by participants using a 1–5-star system. If participants do not like the proposed nudges, they can press the ‘x’ button to receive different suggestions.

Participants will be randomised to one of three versions of the MindCraft app:

#### Personalised digital AI-based recommendations (experimental intervention group)

The experimental arm will deliver personalised behavioural recommendations (AI nudges) (Figure 4a). Personalised nudges are generated using a contextual bandit algorithm, a reinforcement learning method that balances exploring new nudges and exploiting successful ones based on user context and observed rewards. Each nudge is modelled as an action, and the user’s self-reported mental health state constitutes the contextual input. Reward signals are derived from user engagement, specifically nudge completion and the subsequent user-provided rating on a 0–5 scale (Figure 4b). To enhance data efficiency and clinical relevance, a rule-based decision tree first curates a subset of eligible nudges based on the user’s current state, prioritising clinician-endorsed nudges associated with predefined outcomes (e.g., motivation or sleep quality). The contextual bandit algorithm then selects a nudge from this reduced action set, balancing exploration and exploitation. Model parameters are updated using batched user feedback processed via a dedicated learning endpoint, allowing iterative refinement of nudge selection over time.

**Figure 4.**
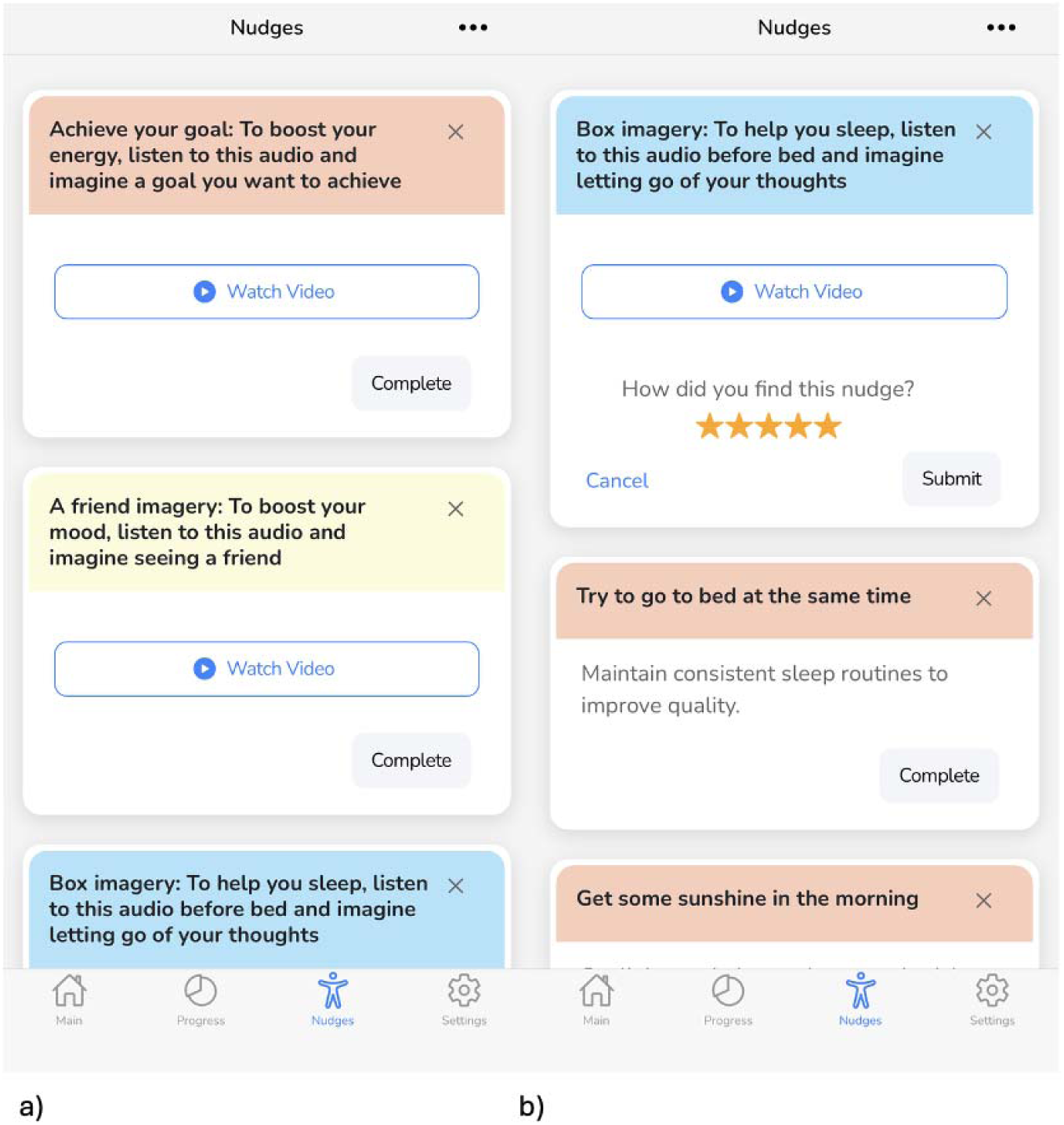
Nudges tab.

#### Non-personalised digital recommendations (active control group)

The active control will be non-personalised digital self-help recommendations using the same content as the personalised AI recommendations, but randomly generated.

#### Self-monitoring only (control group)

The control will consist of self-monitoring via the MindCraft app, including daily ratings and charts of active trackers. No nudges are delivered in this version of the intervention.

### Outcomes

The baseline and 4-week follow-up assessments consist of web-based, self-reported, validated mental health measures.

At baseline, the following screening measures and descriptive data will be collected.

– Demographics: age, gender, ethnicity, education level, location (school)
– Presence of learning difficulties, organic brain disease, or severe neurological impairment that prevents independent use of smartphone app (yes/no)
– The Mood Disorder Questionnaire (MDQ) [25] is a screening instrument for bipolar disorder. The MDQ includes 13 questions plus items assessing symptoms and functional impairment. A total score is calculated where a “Yes” provides a score of 1 and “No” is 0. To meet the threshold for bipolar disorder, respondents need to have a score of 7 or more for questions 1-13, “yes” for symptoms clustered in the same time period and symptoms causing either “moderate” or “serious”.
– The Screening to Brief Intervention (S2BI) is a brief screening tool developed by the National Institute on Drug Abuse (NIDA) to assess for substance use disorder risk among adolescents 12-17 years old [26]. S2BI ask respondents about the frequency of past-year use of tobacco, alcohol, marijuana, unprescribed prescription drugs, illegal drugs, inhalants and herbs/synthetic drugs. According to their responses (“Never”, “Once or twice”, “Monthly”, “Weekly or more”), this tool triages respondents into one of three levels of substance use disorder risk: no reported use, lower risk and higher risk. The questionnaire has been validated in adolescent samples [27].

#### Primary outcome

The primary outcome is the mean change in total difficulties score measured using the Strengths and Difficulties Questionnaire (SDQ) from baseline to 4 weeks between the three experimental, active control and control groups.

The Strengths and Difficulties Questionnaire (SDQ) [28] is a brief behavioural screening questionnaire used in 2-17-year-olds. It consists of 25 items divided between 5 scales: emotional symptoms (5 items), conduct problems (5 items), hyperactivity/inattention (5 items), peer relationship problems (5 items), and prosocial behaviour (5 items). The first 20 items are added together to generate a total difficulties score. In low-risk or general population samples, an alternative three-subscale version of the SDQ can be used, divided into ‘internalising problems’ (emotional + peer symptoms, 10 items), ‘externalising problems’ (conduct + hyperactivity symptoms, 10 items) and the prosocial scale (5 items).

#### Secondary outcomes

The secondary outcomes are the mean change in the following measures from baseline to week 4:

– The Eating Disorder Diagnostic Scale (EDDS) [29] is a 22-item self-report tool designed for individuals aged 13 to 65 that simultaneously assesses diagnostic symptoms of anorexia nervosa, bulimia nervosa, and binge-eating disorder by asking about body image, eating patterns, and compensatory behaviours over the past 3 to 6 months. Items use a combination of Likert, yes-no, frequency, and write-in response formats. An overall eating disorder symptom composite score can be computed by standardising and summing up scores across all items (except for items asking for weight, height, and birth control pill use), and a cut-off score of 16.5 accurately distinguishes clinical patients from healthy controls [30].
– The Sleep Condition Indicator Questionnaire (SCI) [31] is an 8-item rating scale measuring sleep problems. The eight items consist of a general question on the period of time the individual has had sleep problems. These items are rated on a scale from 0 to 4; 4 being the best (e.g., no problem/less minutes to fall asleep/very good sleep quality) and 0 being the worst (e.g. >1 year of sleep problems/more than an hour to fall asleep/very poor sleep quality).
– The Self-Injurious Thoughts and Behaviours Interview (SITBI) [32] is a structured interview that assesses the presence, frequency, and characteristics of a wide range of self-injurious thoughts and behaviours, including suicidal ideation, suicide plans, suicide gestures, suicide attempts, and non-suicidal self-injury (NSSI). Only the NSSI section of the interview will be used, modifying the term NSSI to “self-harm” to include all self-harm behaviour regardless of intent.
– The Self-Efficacy Questionnaire for Children (SEQ-C) [33] includes three 8-item scales that measure academic, social, and emotional self-efficacy. The academic self-efficacy scale includes questions about the person’s perception of achieving academic goals. The social self-efficacy scale addresses social challenges, and the emotional self-efficacy scale includes questions about coping with unpleasant problems or events.
– The World Health Organisation- Five Well-Being Index (WHO-5) [34] is a short self-reported measure of current mental well-being suitable for children aged 9 and above. The WHO-5 consists of five statements, which respondents rate on a scale from 0 to 5 in relation to the past two weeks (0 = “At no time”, 1 = “Some of the time”, 2 = “Less than half of the time”, 3 = “More than half of the time”, 4 = “Most of the time”, 5 = “All of the time”). The total raw score, ranging from 0 to 25, is multiplied by 4 to give the final score, with 0 representing the worst imaginable well-being and 100 representing the best imaginable well-being. Item response theory analyses in studies of CYP indicate that the measure has good construct validity as a unidimensional scale measuring well-being in this population [35].

Additional secondary outcomes include app data and engagement metrics:

– Active trackers: the questionnaire contains 8 compulsory questions (sleep quality, sleep quantity, mood, motivation, anxiety, loneliness, appetite, and exercise) and 11 optional questions (racing thoughts, negative thinking, hopefulness, headaches, irritability, confidence, sociability, energy levels, productivity, self-care, leisure, and a custom metric called “your measure”). Most self-reported questions are on a scale of 1-7 and contain a slider. Sleep quantity and exercise time record the number of hours and minutes completed per day.
– Passive trackers on the app: location, step count, background noise level (Android only), ambient light (Android only), screen brightness (iOS only), battery level, phone usage (Android only), MindCraft app usage. From the raw passive data, distinct features will be engineered [20], for example, total daily steps (n), total distance travelled (m), total ambient light at night (lx), total background noise (dB), total phone usage (min).
– Usability/engagement metrics over the study period for the MindCraft app, including the number of data entries per active tracker, the number of individuals enabling passive tracking, the number of individuals enabling each optional active tracker, and the total number of days/weeks the app was accessed.

### Sample size

A power analysis was conducted based on the minimum clinically important difference (MCID) for the primary outcome, the Strengths and Difficulties Questionnaire (SDQ). Based on the predictive modelling study by Krause et al. [36], the most precise anchor-based MIC for the SDQ was estimated at −1.7 points (95% CI −2.2, −1.2), indicating that an improvement of approximately 8% in SDQ scores may be perceived as clinically important by youths. Variability was estimated from the standard deviation of change scores, which was approximately 5.5, corresponding to a standardised effect size (Cohen’s d) of 0.31.

For a three-arm RCT, a two-sided α = 0.05, and 80% power, a total of 282 participants (94 per group) would be required to detect clinically meaningful between-group differences. Accounting for 60% attrition, which was observed in our external pilot, the recruitment target increases to 705 participants (235 per group). We have implemented changes to the study design and engagement strategies to reduce attrition in the current trial.

It is important to note that the external pilot excluded participants with self-harm or suicidal thoughts (as measured by Item 9 of the Patient Health Questionnaire-9) [37] and therefore included only lower-risk participants who may derive smaller benefits from the intervention. Although the sample size was calculated based on the MCID, pilot sub-analysis indicated that high-risk participants may experience larger intervention effects, suggesting that the study will be adequately powered to detect meaningful changes in this subgroup. As such, the sample size presented here is an estimate and may differ from the actual number required once higher-risk participants are included.

### Statistical Analyses

Where not specified, we will use a Treatment Policy estimand approach, meaning all participants will be included in the analyses according to their randomised allocation.

#### Primary analyses

– Repeated measures analyses (using a mixed effects linear regression model with a random effect on participant) will be used to compare primary outcomes across follow-up points. Timepoint will be included as a categorical variable, and baseline SDQ scores as covariates. The primary endpoint is the intervention difference at 4 weeks. As multiple analyses are performed to compare both the experimental interventions and the active control with the control, and to compare the experimental intervention with the active control, a multiplicity adjustment will be made with a Bonferroni correction. We will also carry out sub-analyses of primary outcomes based on participants’ demographic characteristics/adding them as covariates in the MLM.

#### Secondary analyses

– The same modelling approach as the primary analyses will be used for secondary outcomes.
– A Principal Stratification analysis will be used to estimate the intervention effect, accounting for pre-specified adherence and compliance while retaining the benefits of randomisation. The causal effect among compliers will be estimated using two-stage least squares (2SLS) regression to address potential endogeneity in nudge exposure. Random assignment to treatment will serve as an instrument for actual exposure, with the first stage modelling exposure as a function of assignment and the second stage regressing outcomes on predicted exposure. Robust standard errors will be applied. Mixed-effects linear regression will also be used to analyse repeated measures across follow-up points, including a random effect for participants to account for within-subject correlations. Timepoint will be included as a categorical variable, and baseline SDQ scores as covariates. Multiple comparisons will be addressed using a Bonferroni correction, and sub-analyses will examine outcomes by participant demographics. This approach allows estimation of causal effects among compliers while also assessing changes in outcomes over time.
– Missing outcome data will be handled primarily using the mixed-effects linear regression model, which implicitly accommodates participants with baseline data under the assumption of missing at random (MAR). As a sensitivity analysis, multiple imputation may be performed, incorporating treatment arm, baseline scores, covariates, and additional baseline predictors of outcome or missingness, to assess the robustness of results to the MAR assumption. Additional sensitivity analyses may explore deviations from MAR if required.
– Mediation analyses using structural equation modelling or parametric regression models will be carried out to gain insight into the mechanisms that could explain the potential effect of the interventions on primary outcomes.

### Data Handling

All study data will be collected, stored, and managed in full accordance with the Data Protection Act and the General Data Protection Regulation (GDPR). A document linking participant identities with study IDs will be maintained separately in a secure, password-protected file on the Imperial College London network, accessible only to named members of the research team, and no personally identifiable information will be included in any electronic databases used for statistical analysis. Questionnaire data will be collected using the Qualtrics platform and stored on secure servers compliant with OECD privacy rules and the European Union Directive on Data Protection before being downloaded and stored on Imperial College London’s secure network drives; no hard copies of data will be retained. Smartphone data will be automatically uploaded via encrypted (https) connections to an ISO 27001-compliant digital client server at Imperial College London, stored in dedicated, physically secure, restricted-access facilities, with nightly incremental and weekly full encrypted backups. Identifiable information, including consent forms and primary research data, will be retained for a minimum of 10 years in line with Imperial College London retention policies, with the Principal Investigator acting as custodian. Participants may withdraw from the study at any point prior to data analysis, in which case all associated data will be deleted, unless the participant consents to the continued use of anonymised data collected up to that point. Following the conclusion of the study, anonymised datasets may be shared with other researchers via secure data repositories and disseminated through peer-reviewed publications and conference presentations. All procedures prioritise the confidentiality of participants, with any breach of confidentiality occurring only if disclosure is necessary to protect the participant’s health or safety.

### Patient and Public Involvement and Engagement

A Young People’s Advisory Group (YPAG), comprising 5 young people (4 females and 1 male) with diverse ages, ethnicities, sexual orientations, and experience with digital mental health interventions, was recruited via social media in autumn 2023. The YPAG has contributed to the design and management of the research during the pilot phase, including informing project documentation, recruitment strategies, and app development. The YPAG has been consulted to adapt the study design following the pilot study, and will help to interpret and disseminate the findings. The YPAG will be compensated 25£/hr in accordance with NIHR guidance. Patient and public involvement activities will be reported using the Guidance for Reporting Involvement of Patients and the Public, Version 2- short version.

### Risk Management

There are no foreseeable serious risks associated with this study, but participant well-being and safety will be prioritised throughout. However, potential minor risks include 1) research procedures causing temporary distress, i.e. questionnaires, self-reporting in the MindCraft app, and nudges delivered by the app, and 2) revealing elevated mental health risk. To manage these risks, firstly, participants will be informed of study procedures and will provide fully informed consent, with trained researchers overseeing assessments and clinicians reviewing app content. Specifically, the app includes evidence-based recommendations co-produced by psychiatrists on the research team and YPAG members. Secondly, participants identified at elevated mental health risk during baseline and follow-up assessments, as well as by self-reporting through the app, will be signposted to internal and external mental health support resources, including school mental health and in-app support (Shout, YoungMinds, Best For You NHS, Kooth) and will be offered to be contacted by clinicians on the research team. In the event of disclosure of actual or intended harm to the participant or someone else, parents/guardians and/or the school will be contacted to protect immediate safety, within the limits of participants’ confidentiality. During the research, the participant may suspend using the app at any time. Likewise, the investigator may interrupt the intervention under investigation whenever necessary to preserve the participant’s safety by disabling the nudges.

### Adverse Events

#### Definitions

An Adverse Event (AE) in this trial will be defined, in accordance with ICH-GCP E6 (R2), as any untoward medical occurrence in a participant, whether or not it is considered related to the study intervention. An AE can include any unfavourable and unintended symptom, sign, or other health-related problem temporally associated with participation in the study. Pre-existing conditions that do not change will not be considered AEs, but any subsequent change in severity or new health-related problems will be recorded as AEs.

An AE will be considered serious if it: 1) results in death; 2) is life-threatening (the participant is at risk of death at the time of the event; 3) requires inpatient hospitalisation or prolongs existing hospitalisation; or 4) results in persistent or significant disability/incapacity.

#### Reporting and Classification

All AEs will be recorded by the study team. SAEs will be reported to the Chief Investigator within 24 hours of awareness. Events judged to be both related to study procedures and unexpected will also be reported to the Ethics and Research Governance Coordinator and the Sponsor within 15 days.

#### Severity

AEs will be graded for severity based on clinical judgment. The most severe grade observed during the course of the event will be recorded. Any AE resulting in death will be classified as the highest severity.

#### Causality

All AEs will be assessed for relatedness to the study intervention by a study clinician or designated investigator. Relatedness will be classified as:

– Related: There is a reasonable possibility that the AE is causally related to the study intervention.
– Not related: The AE is more likely to be explained by another cause.

#### Expectedness

AEs will be assessed as expected or unexpected based on the known safety profile of the digital intervention, literature, and prior studies of similar interventions.

#### Outcome

The outcome of each AE will be categorised as resolved/recovered, resolved with sequelae, ongoing, or unknown.

## Discussion

### Expected Outcome

While digital mental health tools show promise for CYP mental health, there is limited evidence on the implementation of AI-enabled tools in real-world settings. This protocol outlines a school-based, three-arm RCT evaluating the effectiveness of personalised AI-informed mobile behavioural interventions delivered via the MindCraft mobile app to support adolescent mental health. This study seeks to generate empirical evidence on the potential use of AI-driven mobile applications as a scalable tool for mental health prevention and early intervention in community CYP, complementing existing mental health support.

### Methodological Considerations

In conducting a digital mental health study within school settings, a range of operational considerations are anticipated. First, recruitment within educational settings necessitates careful communication with school leadership to align the study’s conduct with schools’ existing cultures and demands, minimise disruption to teaching, and ensure transparency in research activities [38,39]. However, recent UK school policies restricting or banning smartphone use during the school day may limit students’ access to personal devices, creating recruitment challenges [40] and influencing outcomes by reducing intervention engagement. This may be mitigated by scheduling data collection and app use outside school hours. Secondly, maintaining participants’ engagement with the app self-reporting over the four-week study period will represent a substantial challenge, as is commonly observed in digital interventions [41,42]. Although passive tracking data may be affected by device and individual variability, it provides objective insights with minimal participant burden, which, together with AI-driven personalised nudges, is expected to enhance engagement and adherence [20]. Finally, study design choices should prioritise representativeness and generalisability by recruiting a diverse participant sample, ensuring that interventions are relevant across varied sociodemographic contexts [43].

Ethical challenges, including informed consent, confidentiality and safeguarding procedures, should also be addressed by research protocols. The study will implement informed written consent procedures, including opt-out parental consent where appropriate, to promote students’ autonomy and inclusivity across diverse socioeconomic backgrounds [21]. Compliance with data protection regulations and transparent reporting practices will be integral to the study’s ethical governance to address ethical concerns for young people and governance preferences for mental health data [44,45,46], including access control, transparency and confidentiality. Comprehensive safeguarding and risk management procedures will be embedded in the conduct of the study, drawing from recent literature and policy guidelines [47,48,49]. We will employ a dedicated safeguarding lead for the study, maintain transparency about safeguarding procedures, respond promptly to mental health needs and report implementation and outcomes qualitatively [47].

## Conclusion

This study has the potential to inform the integration of AI-enabled digital mental health tools within school settings. If successful, personalised nudges could complement existing school-based mental health prevention and early intervention programs, providing a low-cost, scalable approach to support adolescent mental health. Findings may also guide policymakers on the safe deployment of AI technologies in educational contexts, ensuring alignment with safeguarding standards and digital ethics frameworks.

## Data Availability

The datasets generated and analysed during the current study will be available on Open Science Framework (OSF) within 12 months of completion.

https://osf.io/qb8f9/

## Declarations

## Ethics approval and consent to participate

The Study Coordination Centre has obtained approval from the Imperial College London Research Ethics Committee (ICREC ID 7134492) on 28/09/2025. The study will be conducted in accordance with the recommendations for physicians involved in research on human subjects adopted by the 18th World Medical Assembly, Helsinki 1964 and later revisions. Participants over 16 will provide written informed consent. For participants under 16, we will use opt-out consent from parents/guardian consent. Written assent will also be obtained from participants under 16 years old. Imperial College London holds negligent harm and non-negligent harm insurance policies, which apply to this study. Individual participants will not receive direct reimbursement; however, participating schools may, at their discretion, receive the funds centrally to support educational or research-related activities.

A Data Monitoring Committee is not required because this study involves a low-risk digital intervention with no investigational drugs or invasive procedures, and participant activities are limited to app-based self-reporting and passive data collection. Trial oversight, including conduct, data integrity, and participant safety, will be managed by the study investigators through regular internal review, with no interim analyses or stopping guidelines needed due to the minimal risk.

## Consent for publication

Not applicable.

## Availability of data and materials

The datasets generated and analysed during the current study will be available on Open Science Framework (OSF) on https://osf.io/qb8f9/ within 12 months of completion.

## Publication Policy

The PI (principal investigator) and study team have developed a plan to promote the dissemination and implementation of the study findings to stakeholders. Results from this study will be reported and disseminated through peer-reviewed journals, conference presentations, and publications on websites, including the Imperial website and other innovative methods. Anonymised data will be made available for secondary analysis following publication of the main results, approximately 12 months post-publication on OSF. The YPAG (Young People Advisory Group) will guide dissemination and implementation efforts to community audiences

## Authors’ contributions

DN, MDS, and AAF conceptualised the study. BK & AAF created the Mindcraft App platform and its adaptations. AV supported app development and maintained the app software. AF, DN, and MDS gained ethical approval and conducted recruitment and data collection. AF and BK curated the data and performed data analysis. JE provided support for statistical analyses. AF drafted the original manuscript. All authors reviewed and approved the final version of the manuscript for publication.

## Acknowledgments

We acknowledge the members of our YPAG (Amalia Blank, Gigliola Wong, Suhaas Sabbella and Theresa Martins) for their input in the development of this protocol.

